# Control of COVID-19 transmission on an urban university campus during a second wave of the pandemic

**DOI:** 10.1101/2021.02.23.21252319

**Authors:** Davidson H. Hamer, Laura F. White, Helen E. Jenkins, Christopher J. Gill, Hannah N. Landsberg, Catherine Klapperich, Katia Bulekova, Judy Platt, Linette Decarie, Wayne Gilmore, Megan Pilkington, Trevor L. McDowell, Mark A. Faria, Douglas Densmore, Lena Landaverde, Wenrui Li, Tom Rose, Stephen P. Burgay, Candice Miller, Lynn Doucette-Stamm, Kelly Lockard, Kenneth Elmore, Tracy Schroeder, Ann M. Zaia, Eric D. Kolaczyk, Gloria Waters, Robert A. Brown

## Abstract

**Importance:** The coronavirus disease 2019 (COVID-19) pandemic has severely disrupted United States educational institutions. Given potential adverse financial psychosocial effects of campus closures, many institutions developed strategies to reopen campuses in the fall despite the ongoing threat of COVID-19. Many however opted to have limited campus re-opening in order to minimize potential risk of spread of SARS-CoV-2.

**Objective:** To analyze how Boston University (BU) fully reopened its campus in the fall of 2020 and controlled COVID-19 transmission despite worsening transmission in the city of Boston.

**Design:** Multi-faceted intervention case study.

**Setting:** Large urban university campus.

**Interventions:** The BU response included a high-throughput SARS-CoV-2 PCR testing facility with capacity to delivery results in less than 24 hours; routine asymptomatic screening for COVID-19; daily health attestations; compliance monitoring and feedback; robust contact tracing, quarantine and isolation in on campus facilities; face mask use; enhanced hand hygiene; social distancing recommendations; de-densification of classrooms and public places; and enhancement of all building air systems.

**Main Outcomes and Measures:** Between August and December 2020, BU conducted >500,000 COVID-19 tests and identified 719 individuals with COVID-19: 496 (69.0%) students, 11 (1.5%) faculty, and 212 (29.5%) staff. Overall, about 1.8% of the BU community tested positive. Of 837 close contacts traced, 86 (10.3%) tested positive for COVID-19. BU contact tracers identified a source of transmission for 51.5% of cases with 55.7% identifying a source outside of BU. Among infected faculty and staff with a known source of infection, the majority reported a transmission source outside of BU (100% for faculty and 79.8% for staff). A BU source was identified by 59.2% of undergraduate students and 39.8% of graduate students; notably no transmission was traced to a classroom setting.

**Conclusions and Relevance:** BU was successful in containing COVID-19 transmission on campus while minimizing off campus acquisition of COVID-19 from the greater Boston area. A coordinated strategy of testing, contact tracing, isolation and quarantine, with robust management and oversight, can control COVID-19 transmission, even in an urban university setting.

**Key Points:** *Question:* Can a multi-faceted approach lead to control of COVID-19 transmission and spread on an urban campus?

*Findings:* Despite a second wave of SARS-CoV-2 in the greater Boston area, Boston University was able to minimize outbreaks by means of active surveillance of campus populations, isolation of infected individuals, early, effective contact tracing and quarantine, regular communication, excellent data systems, and strong leadership. Most transmission appeared to occur off campus and there was no evidence of classroom transmission.

*Meaning:* Using the main axioms of infection control including frequent testing, vigorous contact tracing, and rapid isolation and quarantine, and a strong leadership structure to ensure nimble decision-making and rapid adaption to emerging data, controlling the transmission and spread of SARS-CoV-2 on an urban campus was feasible despite worsening local transmission during the course of the semester.

## INTRODUCTION

The SARS-CoV-2 global pandemic resulted in nearly 1.8 million deaths worldwide in 2020.^1^ The initial surge of United States cases had a devastating impact on universities and colleges due to widespread campus closures in spring 2020.^2^ Faced with serious financial challenges and adverse social impacts associated with continued closure, some universities developed multi-layered COVID-19 risk mitigation strategies to allow campuses to reopen during the fall semester.^3,4^

Boston University (BU) is a private university with a student/staff/faculty population of ∼40,000 individuals located in the heart of a large US city, a scenario for potential widespread COVID-19 transmission. Despite these challenges, the BU administration pursued an aggressive risk mitigation strategy involving widespread asymptomatic screening for COVID-19, environmental modifications, classroom de-densification, and contact tracing, isolation and quarantine, in order to allow its students to return to in-person learning in the 2020 fall semester. Here we describe the BU experience as a case study offering important lessons that may be broadly applicable to other higher education institutions.

## METHODS

Initial planning, starting in March 2020, centered on active surveillance for asymptomatic and symptomatic cases via on-site molecular testing for SARS-CoV-2 (Supplement). This involved setting up systems for high-throughput laboratory testing, timely communication of results, rapid contact tracing, isolation of infected individuals, and quarantine of close contacts (Figure 1).

**Figure 1.**
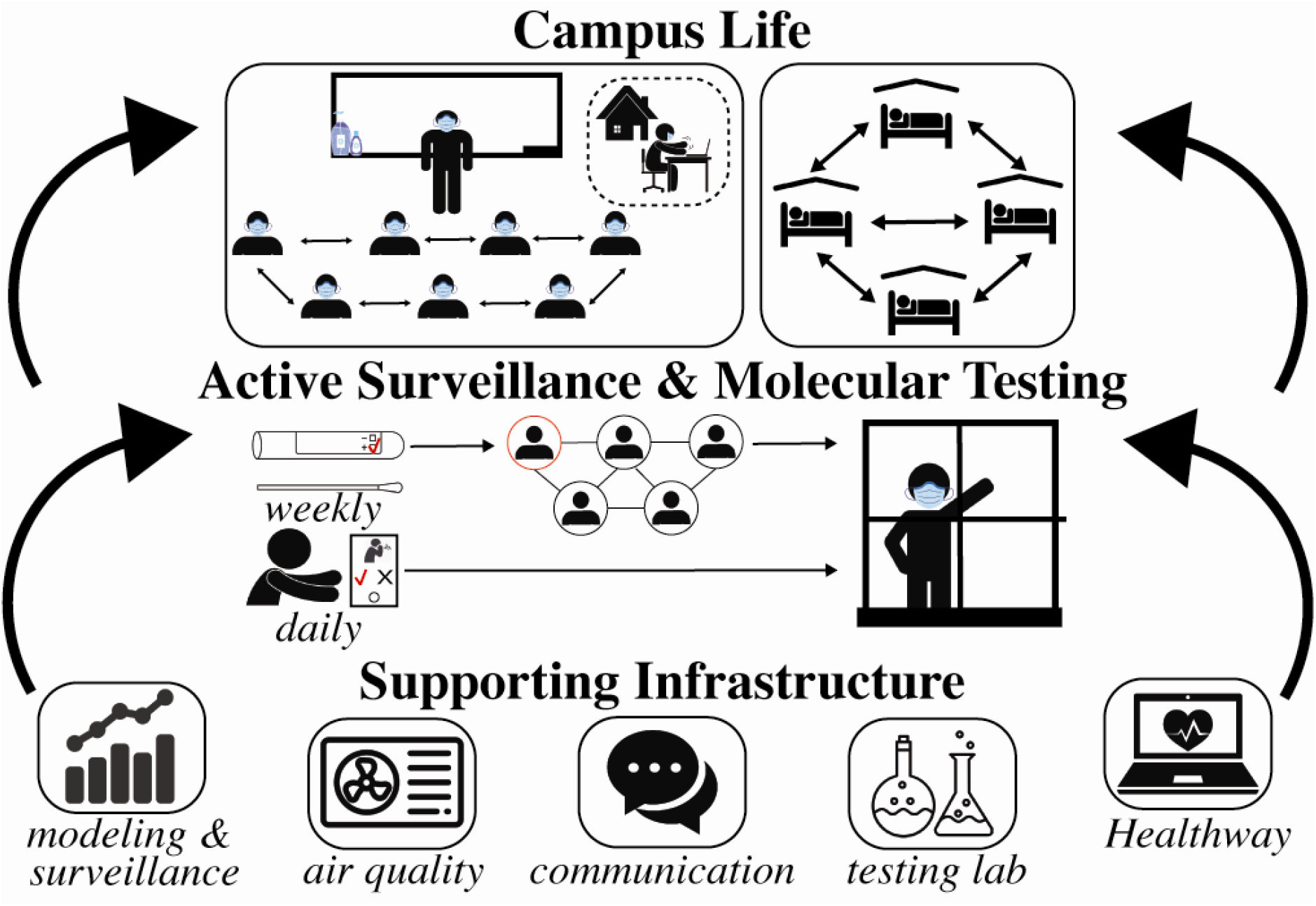
Schematic of the major components of the campus COVID-19 control strategies. (Figure credit: Gina Mantica, Boston University)

Surveillance was complemented by additional control measures, including mask use, enhanced hygiene practices, social distancing recommendations, daily health attestations, de-densification of classrooms and public places, and enhancement of all building air systems. This process was aided by mathematical modeling, multiple data systems, 24 hour/7 day per week monitoring, and data-driven strategy refinements throughout the semester. This required a coordinated leadership and management structure (Figure S1), through interlocking management committees and leadership groups and communication strategies.

### Setting

BU, a large urban university in Boston, has two campuses: the larger main campus (Charles River Campus [CRC]) with over 30,000 members, and a smaller medical campus (BU Medical Campus [BUMC] consisting of the Schools of Dentistry, Graduate Medical Sciences, Medicine, and Public Health) with nearly 10,000 members. In fall 2019, there were 11,439 students living on campus, 23,150 off campus, 10,514 staff and 4,169 faculty (faculty and staff numbers exclude overseas operations).

### Management

Starting in spring 2020, several groups were convened to coordinate COVID-19 control efforts on campus, including monitoring incoming data, modifying campus operations, implementing best public health and medical practices, surveillance, and budgeting (Supplement 1.1, Figure S1).

### Learn from Anywhere (LfA)

BU implemented hybrid teaching in which all undergraduate and graduate students could attend classes in-person or remotely. This allowed classes to simultaneously accommodate on-line only students using Zoom software and in-person students.^5^ Older faculty or those with high-risk medical conditions could opt for on-line only teaching.

### BU SARS-CoV-2 Testing

#### Testing laboratory

BU developed its own polymerase chain reaction (PCR) testing laboratory. Test site staff observed anterior nasal swab sample self-collection by students, faculty, and staff (Supplement 1.2, 1.3).

#### Information Technology

A system was developed by the University’s Information Services and Technology experts linking electronic medical record systems for students, faculty, and staff, and the laboratory information system in the testing facility with the web-based system for daily symptom attestation of all community members on campus, reservations for tests and negative test reporting.

#### SARS-CoV-2 testing categories

Based on guidance from public health authorities, the University developed four SARS-CoV-2 testing categories, reflecting different risks of exposure. These categories determined testing frequency, ranging from twice weekly for category 1 (e.g., on-campus undergraduates) to no testing for category 4 (e.g., students, faculty, or staff entirely off-campus) (Supplement 1.4).

#### Managing and responding to test results

All individuals with a negative test were automatically notified by email to access their test results through a secure website. University health staff directly contacted individuals who tested positive.

### Contact Tracing

The contact tracing protocol was based on CDC and Massachusetts Community Tracing Collaborative processes^6,7^, with adaptation from BU academic programs and student input. Contact tracers followed a detailed script to identify all close contacts, which were defined as someone within six feet of an infected person for 15 minutes or more over a 24-hour period (Supplement 1.5).

### Isolation and Quarantine

Students who tested positive for SARS-CoV-2 had to isolate for 10 days after symptom onset and resolution of fever for at least 24 hours, and with improvement of other symptoms, or for 10 days from the positive test date if asymptomatic.^8^ Students identified as a close contact had to quarantine for 14 days from the exposure date. Based on evolving data from the CDC, this period was reduced to 10 days of quarantine on November 20^th^.^9^ Such students living on campus were moved to special dorms operated by BU (650 units for quarantine and 342 for isolation) (Supplement 1.6-1.8).

### Additional Campus Control Measures

Additional measures including face mask use, enhanced hand hygiene, social distancing recommendations, daily health attestations, de-densification of classrooms and public places, and enhancement of all building air systems including optimization of filtration units were implemented (Supplement 1.9-1.10).

### Mathematical Modeling

We used probabilistic SEIR (susceptible-exposed-infectious-recovered) transmission modeling during summer 2020 to understand the expected relative efficacy of interventions for reducing COVID-19 transmission in the BU community with the goal of achieving only linear increases of cases from transmission outside BU (Supplement 1.11). We used a stochastic agent-based model, implemented using the COVID agent-based simulator (covasim) framework.^10^

### Communications, Surveillance and data management

BU developed a dedicated COVID-19 external communications platform called Back2BU.^11^ The pre-existing ecosystem of data warehousing and analysis systems was augmented to support the data storage, management, and analysis requirements necessary to allow for near-real time surveillance of BU’s COVID-19 response. Surveillance efforts focused on monitoring not only standard metrics around incidence, isolation, and quarantine, but also testing, contact tracing, and compliance with campus control measures. An external dashboard was created and updated daily to allow anyone to track BU metrics (Supplement 1.12-1.15). In addition, an augmented, internal dashboard was created to aid the various groups working with BU leadership to monitor and adapt the BU COVID-19 response.

### Compliance

Compliance with testing and attestation of symptoms were tracked electronically from October. The dean of students collected reports of violations of campus mandates, including gatherings, masking, and physical distancing (Supplement 1.16). Spot checks of compliance were conducted in parallel using trained observers (Supplement 1.17).

### Fall 2020 Move-In

A comprehensive staggered return to campus scheduled was adopted to reduce crowding and lines, giving students the time to test and the university the time to respond to any positive cases on arrival (Supplement 1.18).

### Ethics

The plan for this analysis was reviewed by the Boston University CRC Institutional Review Board (IRB) and was classified as non-human subjects research. The BUMC IRB reviewed the safe behavior quality improvement project (Supplement 1.17) and determined it to be exempt.

## RESULTS

We structure our results to provide information on the operational aspects of the systems BU developed to manage the epidemic and resulting epidemiological features of COVID-19 on the university campus and in the surrounding community. In general, we describe results during the semester: September 3-December 19, 2020; however, we include some results from initialization of systems during the summer.

### Operational Results

Overall, the systems designed in the summer to mitigate the pandemic performed well throughout fall semester.

#### Housing

While 99% (7266) of graduate students lived off-campus, most undergraduates lived in on-campus housing, including a total of 7,131 students as of October 13, 2020, representing 67% of the fall 2020 capacity. Due to de-densification efforts, 3453 (48%) of students lived alone and 3678 (52%) of students lived with one roommate (Supplement 1.5).

#### Testing

Initial operations, staffing and supplies aimed for a maximum of 6200 tests per day (∼42,000 tests per week) and to turn around results within 24 hours of collection. The laboratory reached a stable run rate by October, averaging 5-6,000 tests per day, with lower volume on weekends. Testing turnaround times decreased dramatically over the semester, leveling off to around 12-15 hours between sample collection to receipt of results (Figure 2A). The BU testing laboratory conducted 467,382 tests during the fall semester (517,357 including the pre-semester move-in).

**Figure 2.**
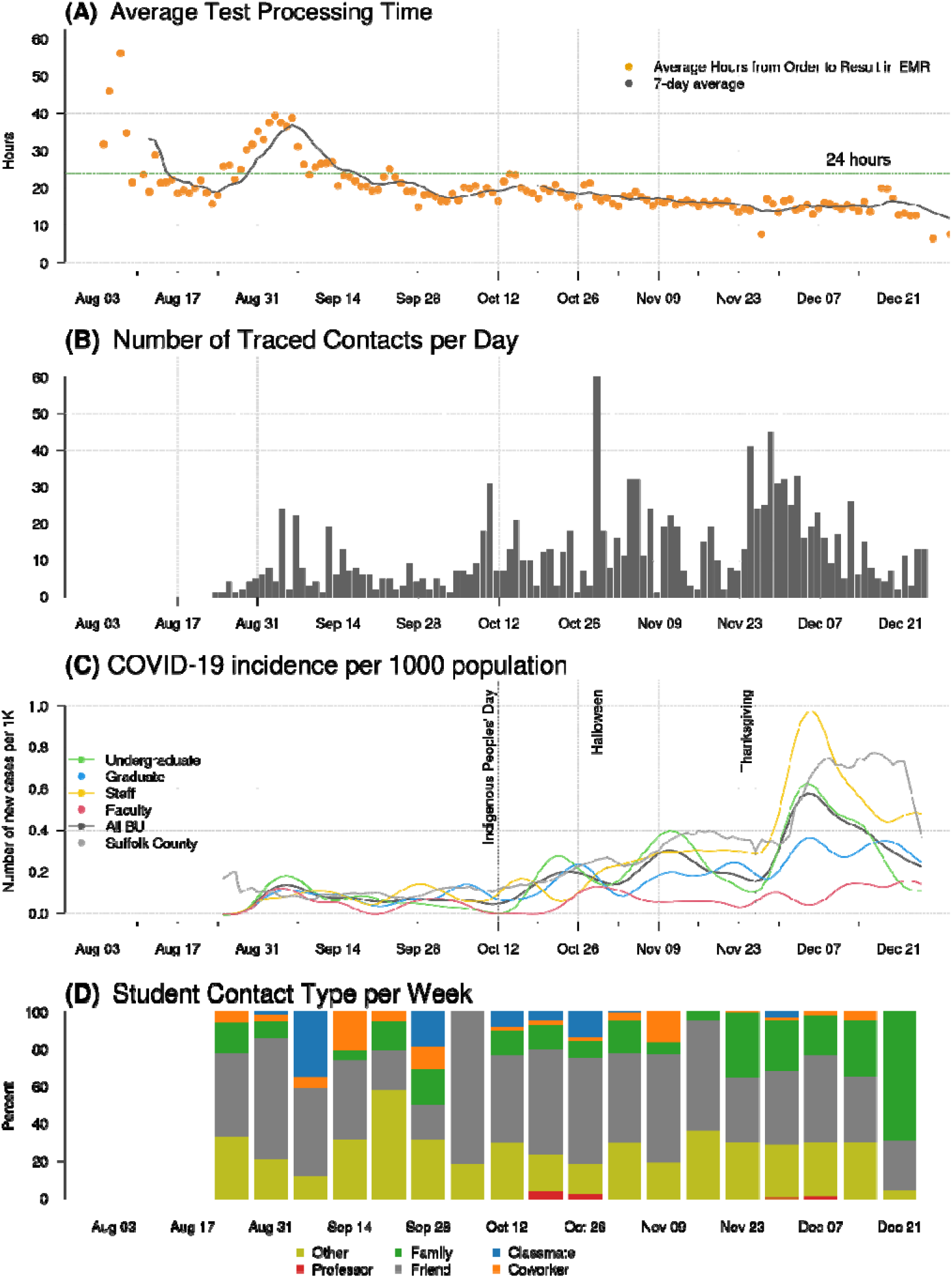
A) Average daily testing turnaround time. B) The number of close contacts traced by week. C) BU COVID and Suffolk county cases per 1000 from August 25, 2020 to December 24, 2020 with a seven day smoother applied. Cases are shown overall, and by key subpopulations, as noted in the legend. D) For students who tested positive, the affiliation of their close contacts: classmate, faculty, coworker, friend, family, and other.

#### Contact Tracing, Quarantine and Isolation

On average within 6.4 hours (median: 3.0, range: 0-42 hours) after receipt of a positive test, close contacts of a positive BU case were notified of their exposure. However, this average conceals improvements over time, falling from ∼10.1 hours in September to 5.4 in December 2020. Students and employees (faculty and staff) identified 3.1 and 0.5 BU close contacts per case on average, respectively. The number of individuals needing to be traced increased with increasing case numbers throughout the semester (Figure 2B).

Despite having quarantine and isolation capacities of 650 and 342 respectively, occupancy only reached a maximum of 13.7% and 12.9% respectively at any one time (Figure S3).

#### Compliance

On average 12% of on-campus students were non-compliant with testing or attestation protocols in October and November, lower than off campus students during the same time (51.5% for off campus students in categories 1, 2 and 3 or 20% if including students in category 4) (Figure 3A). Reported violations were most common as the semester began, but gradually tapered off (Figure 3B). Early semester violations were largely off-campus gatherings, which reduced sharply by November. Compliance with face masks was high throughout the semester.

**Figure 3.**
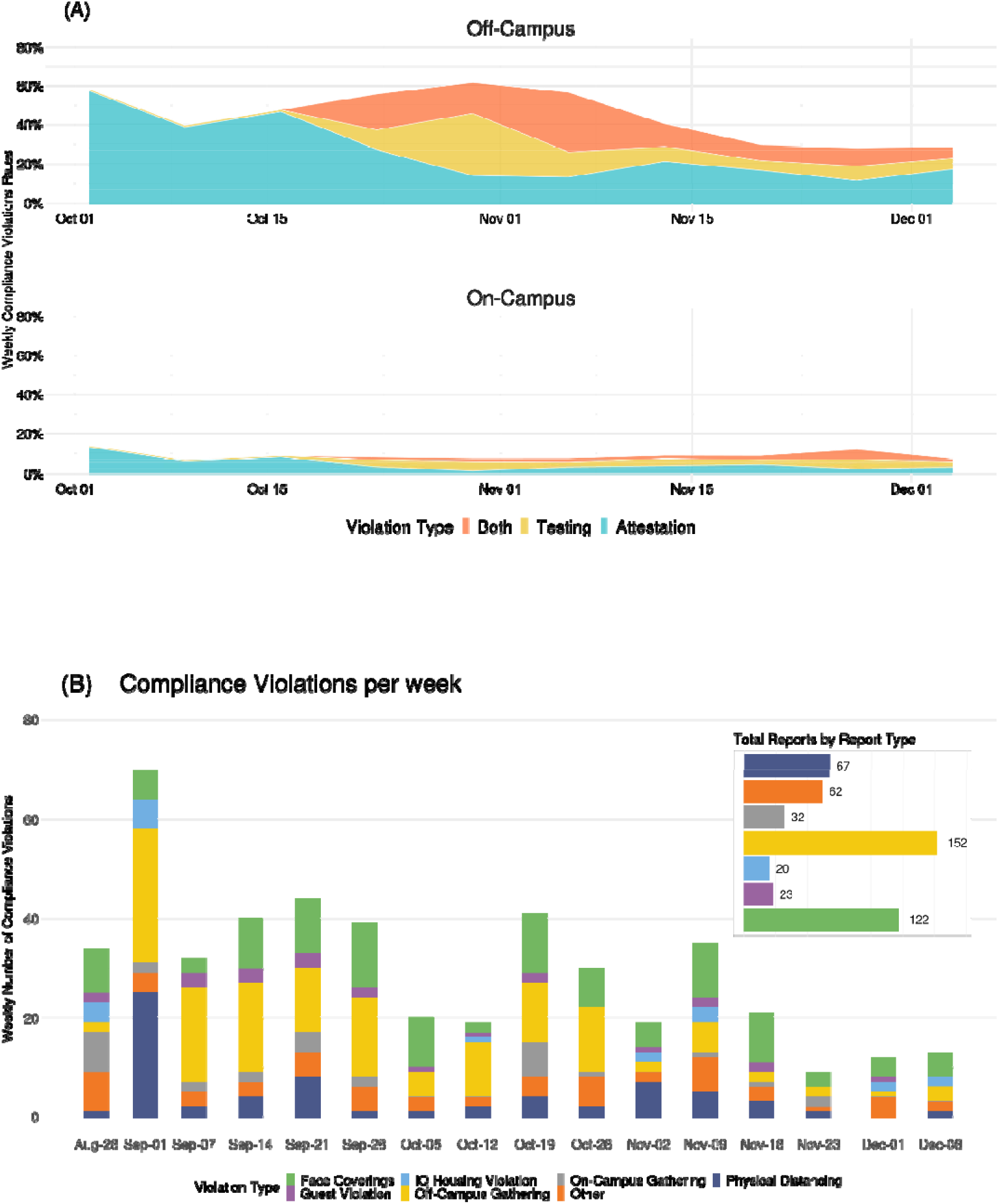
**A)** Weekly percentage of off-campus (top) and on-campus (bottom) students in violation of testing and/or attestation requirements. Off campus compliance only considers students in categories 1-3, who had a testing and attestation requirement. **B)** Total weekly number of reported compliance violations for control measures other than testing or attestation, by type of violation. (Inset: Total number of violations by type over the Fall semester.)

#### Classroom Density

Class attendance data were not collected systematically. An upper bound on attendance levels is given by the percentage of registered students indicating through LfA that they would attend in person. For medium, large, and “very large” classes, this number was on average approximately 45% in October (with medians similar) but dropped to roughly 25-30% in November. For small classes, while the mean was similar to the other groups, the variability was substantially greater (Figure 4).

**Figure 4.**
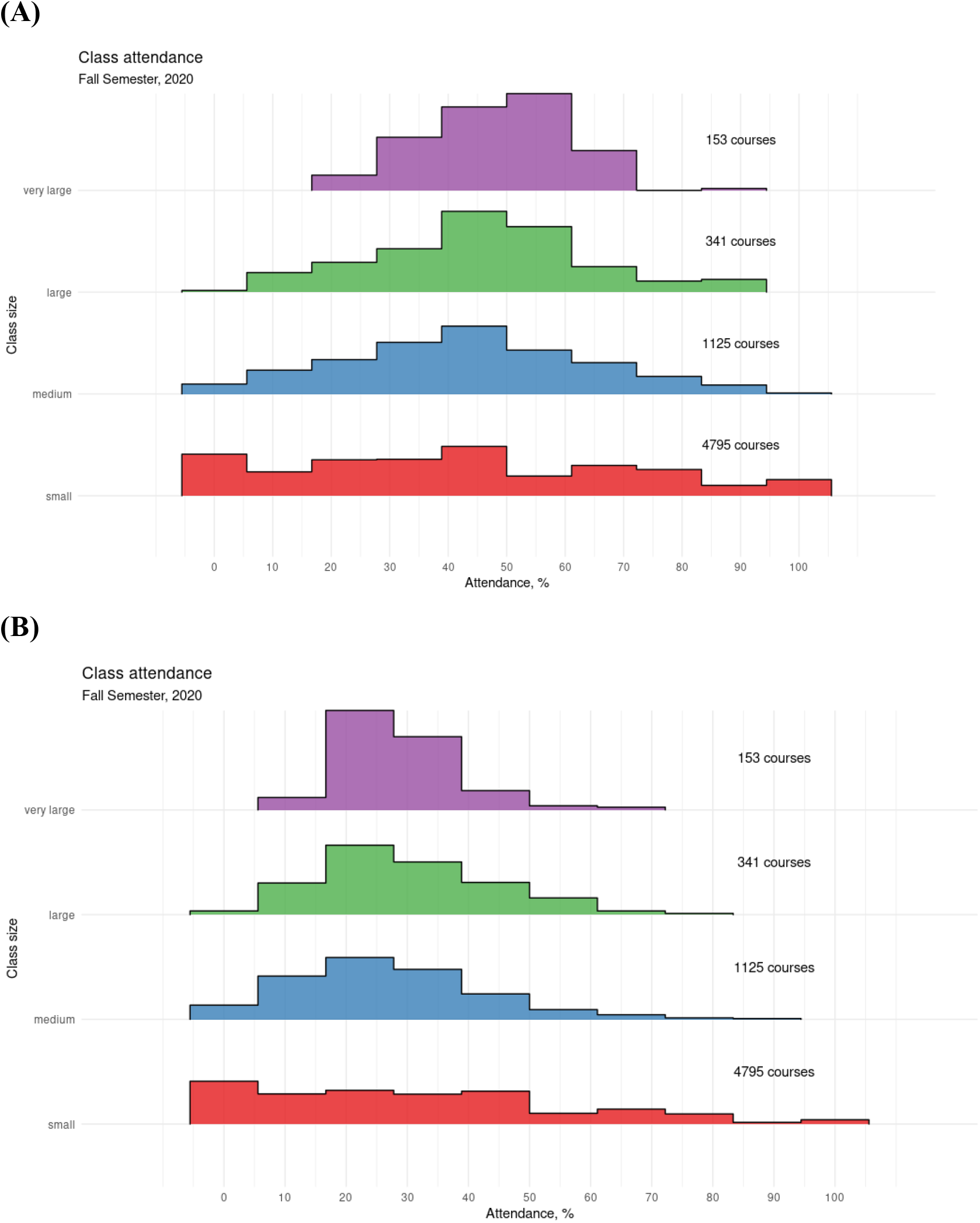
Percentage of registered students indicating through LfA that they would attend class in person, over all BU classes, by size of class, for October (A) and November (B). Class size is based on total number of registered students: small (3 to 25); medium (26 to 50); large (51 to 100); and “very large” (greater than 100).

### Epidemiology of COVID-19 at BU and in Boston

#### Changing epidemiology of COVID-19 in Boston

Following statewide control measures in spring 2020, case numbers in Suffolk County (which includes Boston) averaged approximately 0.1 case per 1000 population through mid-October. As state restrictions eased and the weather became colder (and people spent more time indoors), cases increased to 0.4/1000 by early November, peaking at 0.75/1000 following Thanksgiving gatherings (Figure 2C). These increasing trends were observed statewide and also in collection of SARS-CoV-2 from Boston area wastewater, indicating that they were not just due to increased testing in the Boston area.^13,14^

#### COVID-19 Burden and Patterns at BU

During the fall semester, 719 individuals tested positive for COVID-19 in the BU community including 496 (69.0%) students, 11 (1.5%) faculty, and 212 (29.5%) staff (Table 1). Cases increased following holidays, particularly Thanksgiving, and concentrated among undergraduate students and non-faculty staff. While the incidence rate among students and staff tracked that of Suffolk County (Figure 2C), there were two distinctions. First, BU surveillance testing detected many asymptomatic cases (37.7% of our total cases), whereas testing elsewhere in Massachusetts was passive and driven by emergent symptoms. Secondly, particularly after Thanksgiving, the BU increase in cases preceded that of Suffolk county. This is most likely attributable to BU’s vigorous testing regime.

**Table 1.** Number of Boston University individuals testing positive for COVID-19 during the fall semester.

|  | Total, N (%) | Students (n=496) |  | Employees (n=223) |  |
| --- | --- | --- | --- | --- | --- |
|  |  | Undergrad, n (%) | Graduate, n (%) | Staff, n (%) | Faculty, n (%) |
| N (%) | 719 (100.0) | 315 (43.8) | 181 (25.2) | 212 (29.5) | 11 (1.5) |
| Asymptomatic, n (%) * | 271 (37.7) | 113 (35.9) | 51 (28.2) | 104 (49.1) | 3 (27.3) |
| Exposure, n (%) |  |  |  |  |  |
| BU HHCC | 6 (0.8) | 3 (1.0) | 2 (1.1) | 1 (0.5) | 0 (0.0) |
| BU CC | 158 (22.0) | 105 (33.3) | 37 (20.4) | 16 (7.6) | 0 (0.0) |
| Non-BU HHCC | 62 (8.6) | 13 (4.1) | 15 (8.3) | 33 (15.6) | 1 (9.1) |
| Non-BU CC | 84 (11.7) | 31 (9.8) | 28 (15.5) | 22 (10.4) | 3 (27.3) |
| Outside BU exposure** | 44 (6.1) | 22 (7.0) | 13 (7.2) | 8 (3.8) | 1 (9.1) |
| Travel^ | 16 (2.2) | 9 (2.9) | 3 (1.7) | 4 (1.9) | 0 (0.0) |
| Unknown | 349 (48.5) | 132 (41.9) | 83 (45.9) | 128 (60.4) | 6 (54.6) |
| Known Exposure, n (%) | 370 (51.5) | 183 (58.1) | 98 (54.1) | 84 (39.6) | 5 (45.5) |
| BU Exposure (HHCC + CC), n (%) | 164 (44.3) | 108 (59.0) | 39 (39.8) | 17 (20.2) | 0 (0.0) |
| Non-BU Exposure (HHCC + CC), n (%) | 206 (55.7) | 75 (41.0) | 59 (60.2) | 67 (79.8) | 5 (100.0) |

\* 2 cases refused to provide data on symptoms, reducing sample size to N=717 for the Asymptomatic variable
\*\*Outside BU Exposure includes cases that have not been on campus and have no known close contact, or recent engagement on campus or with BU community members.
^Travel indicates case returned from out-of-state travel via public transportation (plane, bus, train) 48 hours prior to symptom onset or positive test.
HHCC: household close contact
CC: close contact

Of the students who were close contacts and entered quarantine, 10.3% (86/837) tested positive for SARS-CoV-2 while in quarantine. The test positivity rate for those who self-attested having symptoms was notably higher (4.9%) than that of asymptomatic individuals (0.10%) indicating utility of this feature.

#### Sources and locations of transmission

BU contact tracers identified a transmission source for 51.5% of cases with 55.7% identifying a source outside BU (Table 1). Among infected faculty and staff with a known source of infection, the overwhelming majority reported a transmission source outside of BU (100% for faculty and 79.8% for staff). Students identified more BU contacts as infection sources (39.8% for graduate students and 59.2% undergraduate students, Table 1). Notably, BU household contacts were identified as a source of infection less than 1% of the time (Table 1) and anecdotally, there were no sustained transmission events in on-campus housing, indicative of the efficacy of efforts to control spread in campus housing. When asked to identify close contacts, students rarely identified classmates. Instead, close contacts tended to be friends and, after Thanksgiving, family (Figure 2D). This indicates that while BU students tended to be more likely to identify another BU affiliate as a source of infection, the contacts leading to infection occurred in places out of reach of BU’s interventions.

## DISCUSSION

Although many US higher education institutions reopened in fall 2020, some experienced large COVID-19 outbreaks forcing a return to on-line only education.^15–17^ The BU experience is important because it has an urban campus in a community that experienced high and increasing COVID-19 incidence from August to December 2020 with no option for the university to isolate from the wider community. Despite this, BU benefited from substantial resources including funding to establish and run a SARS-CoV-2 testing laboratory and sampling centers, a hybrid learning approach (LfA), and diverse expertise among university employees in medical epidemiology, modeling, biostatistics, and public health control measures. In addition, strong central leadership; internal communication to students, staff, and faculty; frequent and adaptive testing of students^18,19^; short testing turnaround time; highly effective contact tracing coupled with isolation and quarantine; and vigorous enforcement combined to prevent widespread campus outbreaks of COVID-19 despite the worsening local situation.

Strong leadership structure with multiple subcommittees targeting important aspects of the response supported the interventions. Frequent communication and coordination between these groups ensured that, if a cluster of cases was emerging, all parties were aware. Thus, testing cadences could be adapted, compliance efforts modified, and messaging adapted to blunt any outbreak risk. This coordinated effort was key to ensuring a high level of compliance and the success of planned interventions.

Short turnaround time of results followed by rapid isolation of infected individuals, contact tracing and quarantine of close contacts resulted in limited transmission in the BU community. Faculty and staff were almost always infected outside of the university campus. While most students with a known source of infection reported another BU affiliate as their contact, these infectious events appeared to occur outside of BU housing and instructional settings, where interventions were targeted. When case clusters appeared in settings the university could not directly target, e.g., social or other off campus gatherings, these were quickly controlled, due to our vigorous testing regime, rapid contact tracing, and strict enforcement measures.

Consequently, no major outbreaks were observed and the resulting numbers of cases throughout the semester were consistent with our goal of maintaining a linear, rather than exponential, increase in cases, which was manageable with our intervention strategies.

Surveillance testing facilitated identification and isolation of many close contacts ahead of contact tracing efforts. Importantly, due to the surveillance testing system, BU tended to detect increases in cases before the surrounding community where people were mostly only tested once symptomatic. This is supported by the fact that 37.7% of positive individuals were asymptomatic at the time of isolation indicating that BU was detecting cases early in the disease course. This finding contradicts recently published research suggesting that BU cases led the surrounding area in a causal manner.^20^ Limitations to the latter study include the inclusion criteria resulting in only one Boston-area institution in the analysis and assuming that community transmission arose from the university campus rather than the converse.

BU’s success is consistent with current understanding of best COVID-19 control practices. These strategies, aggressive testing, contact tracing, and quarantine and isolation, have been successfully implemented in many countries, including Singapore, South Korea, and Taiwan.^21–23^ However, unlike these countries, the BU setting did not allow restriction of travel between the campus and nearby community, making this a strong demonstration of the utility of these approaches despite substantial importation of cases from the surrounding community. This can potentially serve as a model for other institutions nested within a broader community.

There were some challenges. Sometimes the contact tracing team was unable to identify contacts due to students’ reluctance to divulge information regarding where they had been or who they had been with. In these cases, coordination between the dean of students and the contact tracing team was critical in identifying other students who were associated with the infected student(s) through team or club membership, so increased frequency of testing (adaptive testing) could be performed.

BU’s approach carries a high financial cost^24^; BU had to implement budget adjustments, including hiring freezes, salary freezes, and several other cost-cutting measures, to meet the cost of these supplemental services and respond to declining revenue because of pandemic-related changes to operations. Ultimately the university was able to meet its financial obligations and avoid large layoffs or other consequential financial impacts. Also, providing students an in-person option was beneficial for varying learning styles and meeting immigration requirements. The remote option also benefitted students who could not attend in person, either due to health considerations or travel restrictions.

BU benefited from being a large research university with much of the required expertise for our strategy available within the university, saving money and facilitating substantial control over the operations. This is clearly not feasible for all higher education institutions. This implies that broader efforts in the community, supported by government public health agencies, are required to control spread. In summary, a multipronged response effectively controlled SARS-CoV-2 spread on an urban university campus, despite rising community burden of disease.

## Data Availability

A limited data set can be made available upon request if accompanied by an acceptable data analysis plan.

## Acknowledgments

We would like to thank the members of the Augmented Budget Committee, Medical Advisory Group, Community Health Oversight Group, Healthway team, and all Boston University faculty, staff, and students who played important roles in keeping the campus safe and healthy during the 2020 fall semester.

