## Supplemental materials for "Control of COVID-19 transmission on an urban university campus during a second wave of the pandemic"

### 1. Program background

#### 1.1 Management

Starting in the spring of 2020, several different groups were organized in order to coordinate all aspects of COVID-19 control on campus. These included an Augmented Budget Committee (ABC), which was responsible for high level coordination of the campus pandemic response with program leads for budget and finance, human resources, residential systems, graduate professional programs, research and clinical operations, libraries, undergraduate education, public health protocols and testing, and external and internal communications. The ABC worked closely with the Contingency Planning Committee and the Medical Advisory Group (MAG) as well as multiple working groups focused on different campus populations (Figure S1). The MAG was responsible for using guiding principles from national, state, and/or local authorities to aid in the development of policies and protocols for the systemic integration of students and employees back to the physical campus. These included determining medical requirements for the safe return of employees and students to campus and development of protocols to support the overall wellbeing of the community campus reopened and during the fall semester. The Community Health Oversight Group (CHOG) was responsible for the daily review of internal and external dashboards that tracked the progress of testing, isolation, contact tracing, quarantine, and compliance of students, staff and faculty to testing and health attestation requirements. The BU Response and Surveillance Team (BURST) was responsible for modeling and analyses of COVID-19 data generated during the implementation of the multi-faceted program described below.

The Auxiliary Services senior leadership team, using the BU MAG and public health guidance, identified 651 single quarantine and 342 isolation beds. Auxiliary Services established the number of COVID-19 quarantine and isolation (Q/I) beds utilizing sophisticated demand and surge models developed by BU researchers (BURST).  The Events and Conferences unit was tasked with managing the non-medical needs of the University's COVID-19 Q/I housing.

#### 1.2 BU COVID-19 testing laboratory

The Massachusetts Department of Public Health (DPH) had approved a CLIA-certified laboratory on the BU Medical Campus to perform SARS-COV-2 testing by reverse transcriptase polymerase chain reaction (rt-PCR). However, in order to have an affiliated laboratory facility on the main campus, the laboratory had to have a license to operate in a new location in compliance with emergency Food and Drug Administration (FDA) guidelines. From March to July, staff were trained and biosafety hoods, robots for pipetting and RNA extraction, and PCR set up, and PCR instruments were installed and tested. Information Services and Technology (IS&T) implemented a new employee EMR and enhanced the existing student EMR to support activities for daily attestations, testing compliance, test scheduling, result reporting, contact tracing, and clearance for work notifications. Concurrently, IST&T projects were implemented to allow automation of laboratory processes and delivery of results automatically to the employee and student EMR systems. The laboratory was designed to operate with two eight hour shifts daily, seven days a week. After submitting an FDA Emergency Use Authorization application and associated data for the laboratory, BU began using a laboratory developed quantitative reverse transcriptase PCR (RT-PCR) assay in July. Similar to the US CDC’s assay, BU used a three target RT-PCR assay targeting the N1 and N2 genetic sequences of SARS-CoV-2 and an RnaseP sequence indicating the presence of human cellular material. While a quantitative assay, positive results were defined if a test delivered an N1 and/or N2 positive result and a positive RnaseP result at a cycle threshold less than 40.

Test site staff observed sample collection by students, faculty, and staff. Self-collected anterior nares swabs were placed into tubes containing one mL of sterile saline solution in a bar-coded tube that was linked to each individual by swiping their BU identification badge just prior to collection. Samples were transported to the laboratory on a daily schedule by a specialized medical courier service, and sampling sites were open seven days a week. This system was stress-tested during a pilot evaluation in July 2020, followed by full-scale implementation beginning in August as students returned to campus.

#### 1.3 Sample collection sites

In June, sample collection sites were identified and modified with required air handling systems to reduce potential transmission at testing sites. The testing staff were separated from those being tested by a plexiglass booth and universal masking. Typically, the process of linking the bar-code on the sample vial to the person and collecting the sample was completed in less than 5 minutes. Moreover, advance registration was required for a testing slot, thereby minimizing crowds and waiting times. After designing policies and procedures, staff were hired and trained. Four sites were created for asymptomatic screening including three on the main campus and one on the medical campus. In addition, there were sites for testing symptomatic individuals on the main campus and for remote testing of students in quarantine.

#### 1.4 Testing categories

Testing category 1 included all undergraduate students; all graduate students living in undergraduate residential housing, BU Academy (a private high school operated by Boston University) cross-registered students taking classes with residential students, and faculty and staff who would be on campus on a regular basis including those who planned to use public transportation to reach campus. Testing category 2 included commuting students residing off-campus attending in-person classes, but with little contact with undergraduate students. Undergraduate students living on or off campus were required to be tested twice a week as were graduate students living on campus; all other categories of individuals in categories 1 and 2 were tested once a week. Category 3 included commuting employees whose job duties required very limited contact with students and who could control their contact with other employees so as to limit interactions to small groups of individuals with appropriate work environment protocols in place and minimal contact hours; and graduate students in degree programs that meet less than once per week. This group was tested before the start of classes and once a week if they planned to be on campus. Testing category 4 was comprised of students, faculty, and staff engaging only in virtual learning, working, and other activities and events and who did not commute to campus. This group was not required to have routine testing performed. Anyone who became symptomatic was advised to have a test done at a symptomatic sample collection center.

#### 1.5 Contact tracing procedures

BU deliberately chose to hire contact tracers from the current BU student community, BU alumni, and other people with similarly good knowledge of the BU campuses and activities so that this knowledge could help them with the job of contact tracing. Contact tracers follow a detailed script to obtain as much information about the potential source of exposure and to identify potential close contacts. Contact tracers were able to establish a rapport with the students that they contacted; this enabled effective case investigation dating 7-10 days prior to symptom onset or positive test date if asymptomatic. This information was helpful to understand where the individual was potentially exposed. To identify who the case had potentially exposed, the contact tracers reviewed individuals that had been in contact with the case from 2 days prior to symptom onset or positive test date if asymptomatic. In addition, the individual was asked where they thought they had most likely been exposed. BU used the CDC’s definition of a close contact: someone who was within six feet of an infected person for a cumulative total of 15 minutes or more over a 24-hour period.^1^

In addition to contacting infected students by cell phone, BU used other contact tracing tools as well. The university’s IS&T department developed a custom PowerBI look-up interface. This in conjunction with information from the Student Health medical record system, helped contact tracers navigate questions, potential contacts, and provided a more informed view to identify potential clusters. Both the student and employee electronic medical records had contact tracing software modules to link cases with contacts. Using analog maps in conjunction with all of these tools and information of the campus assisted in identifying dormitory and work exposures. The contact tracing database was also used to search an individuals’ classmates, teammates, on-campus roommates, and instructors/faculty or a faculty member’s students, which was used to cross check for potential classroom transmission. This helped to identify potential clusters. The student and employee electronic medical records had contact tracing software modules to link cases with contacts. In turn, the university used information found during the case investigation and within the database to inform adaptive testing (increased frequency of testing in suspected clusters of cases). If an individual did not meet criteria to be a close contact, but was part of a social group (e.g., Greek-life membership) or affiliation (e.g., sports team, music group) of recent cases, they were notified of their potential risk of COVID-19 exposure, reminded to follow public health guidelines, and required to increase testing frequency to 3 times per week.

#### 1.6 Isolation

Students living on campus were moved to one of four isolation buildings on campus with a total of 342 units. They were provided with amenities and food deliveries throughout the time period. Students living off campus were offered and advised to move on campus for their isolation period to mitigate the transmission risk to their off-campus roommates.

All isolating students were checked daily via a message from the electronic medical record system to their patient portal and called daily by a nurse. If a student in on-campus isolation was symptomatic, they were also evaluated in-person by an isolation nurse. Telehealth appointments with a clinician were available to students through Student Health Services.

BU employees (faculty and staff) who tested positive had to isolate for the same timeline and were referred to their primary care providers for follow-up. Nurses called to check in every 5 days throughout their isolation period. Due to a work restriction, they were not allowed to come to campus until the full isolation period had passed.

#### 1.7 Quarantine

Students living on campus were moved to one of two quarantine buildings (650 total units) and were provided with their own bedroom, bathroom, amenities and food deliveries throughout the time period. Students living off campus were counseled on quarantine guidelines and best practices. All students in quarantine were checked daily via a message from the electronic medical record to their patient portal. Additionally, a contract tracer called them every 2-3 days and, if symptomatic, a nurse would call daily.

PCR testing was provided for all students in quarantine every 3 days. Students living on campus had remote testing appointments scheduled and were delivered sample collection kits. During a video appointment with a staff member, observed self-collection of an anterior nares sample was performed. Off campus students scheduled an appointment at a separate testing site for symptomatic individuals and those identified as close contacts.

If an off-campus student tested positive, they followed the isolation protocol. If the student continued to test negative, they could be removed from quarantine on day 10 if asymptomatic or on day 14 if symptomatic.

Employees identified as a close contact from a workplace exposure or community exposure were required to quarantine for the same timeline. They were scheduled to have a test at the Health Services Annex, a testing site that was exclusively for close contacts and symptomatic individuals, every 5-7 days throughout their quarantine period. A work restriction was established, and they were not allowed to come to campus, except for testing.

#### 1.8 Campus housing and dining

BU Housing reduced density by removing quads and triples from shared sleeping spaces. Housing assigned students to doubles and singles only, and ResLife created small households to manage shared common areas. An extended fall move-in schedule intentionally limited the number of student arrivals on any given day and required students to select an arrival date and time.

Residence Life (ResLife) assumed responsibility for coordinating Q/I assignments working with Healthway (section 1.13) medical professionals and contact tracers. Students were assigned to one of two quarantine housing buildings based on proximity from their dormitories so that they could move themselves into and out of quarantine. Isolation housing assignments on the Fenway Campus (a location a short distance from the main campus) had a transportation option for students, given the distance involved. Arrivals and departures utilized a PowerBI dashboard developed to track arrivals and departures specifically to manage inventory turnover.

Q/I rooms were pre-stocked with a 10 to 14 day supply of non-perishable foods and beverages. Rooms were provided with linen and bedding, in addition to various self-care necessities a student would need while in quarantine or isolation. BU Dining Services delivered fresh (non-perishable) meals and beverages four times a week. Certified nutritionists from Sargent College reviewed the menus and collaborated with BU Dining Services to support students who had allergies and dietary restrictions with customized meals.

The BU Dining Services team had to reinvent their operations to provide a balanced yet robust, safe dine-in and take-out meal program. Massachusetts public health guidelines required reduced dine-in seating options and dining hall capacities, and new routines were developed around meal preparations, disposable service, and sanitation procedures. The University required weekly testing and daily health screening for all dining staff employees. The BU Dining services culinary team worked closely with nutritionists from our Sargent College of Health and Rehabilitation Sciences on a new six-week menu cycle to ensure students had options to meet every dietary preference.

BU housing stock as of summer 2020 consisted of BU dorms, suites and apartments with single (2930), double (7027), triple (1108) and quad (240) occupancy. De-densification of housing led to 3453 (48%) students living alone and 3678 (52%) with one roommate.

#### 1.9 Additional control measures

Face mask use was mandatory on campus including in classrooms and other university facilities, and outdoor university public places. The only exceptions were for faculty when alone in their own office, and for on-campus students when in their dorm rooms. Enhanced hand hygiene with alcohol-based hand wash was available throughout campus. Students were forbidden from congregating in others’ dorm rooms. University cafeterias provided all meals in sealed containers and the students took their meals away to either eat outside or in their dorm rooms. Lastly, recommendations for social distancing, daily health attestations, de-densification of classrooms and public places such as cafeterias, and enhancement of all building air systems including optimization of filtration units were implemented.

#### 1.10 Daily Health Attestations

All students, faculty, and staff with a sufficient level of on-campus presence (Categories 1 and 2) had to complete a daily electronic health attestation, consisting of screening questions for common COVID-19 symptoms; this was done through a web-based application linked to the appropriate EMR. Students, faculty and staff were cleared to be on campus if they had done their daily attestation, completed SARS-CoV-2 testing in the previous week according to their category requirements, and scheduled their next test.

#### 1.11 Mathematical modelling

We used probabilistic SEIR (susceptible-exposed-infectious-recovered) transmission modeling to determine if the cumulative impact of these interventions could reduce transmission within our community such that most cases would be attributable to exogenous sources (i.e., transmission in from outside the BU community rather than within-BU transmission), leading to linear growth in case numbers over time that is proportional to the assumed constant exogenous rate. Our work included a stochastic agent-based model, implemented using the COVID agent-based simulator (Covasim) framework.^2^ We used institutional data to build classroom and housing networks of interactions at the level of individual students, faculty, and staff, treating all other sources as exogenous sources of infection. We layered on intervention strategies, including frequent testing, contact tracing, and de-densification of housing and classrooms, and projected the relative efficacy of these interventions in reducing case burden under various proposed strategies. Our planning scenarios in the model indicated that the cumulative benefit of the intervention strategies that were ultimately adopted were expected to decrease case growth to be approximately linear over time (Figure S2) under a constant exogenous rate. Details on the model and code are available on GitHub.^3^

#### 1.12 Surveillance and data management

Multiple groups within the University’s Information Services & Technology (IS&T) Department provided coordinated support for modeling and data management activities. The Reporting and Data Architecture group created and completed several automated data collection and database development initiatives; pulling and consolidating information from a variety of key source systems (e.g., Student Registration and Course Scheduling, Student Health, Housing, and ERP and Human Resource systems, etc.) to support reporting and analysis efforts at a wide and more flexible scale. The IS&T Analytical Services & Institutional Research (AS&IR) group supported the University’s analytical and reporting needs. AS&IR worked across a variety of units and departments to identify, consolidate, and feed source data for analysis, as well as map these fields within their source systems for report development and automation of data feeds. In addition, the group developed a suite of internal and public dynamic reports supporting operational processes, monitoring, and data access. Finally, the team provided analytical support to the various working groups and senior leadership as they worked to develop and implement solutions and improvements to propel forward the University’s collective efforts to operate safely and efficiently. The Research Computing Services (RCS) group facilitated engagement between BU faculty and other groups within IS&T, provided programming, infrastructure, and dissemination support for mathematical modeling analysis and reporting, and closely coordinated with the IS&T groups above on data engineering, management, and automation activities.

#### 1.13 Healthway

Healthway was a central organizational structure that involved the integration of add healthcare professionals providing triage and case management, contact tracing, Information Services and Technology (IS&T) systems, result reporting and daily symptom attestation. The Healthway team had access to MAVEN (Massachusetts Virtual Epidemiologic Network) to enter cases and close contacts. Close communication with the Massachusetts Department of Public Health and Boston Public Health Commission was integral to the contact tracing program.

As IS&T functions were developed, including compliance rules, staff members were hired and trained to support the phone responses required to manage the symptom attestations, report positive results, investigate close contacts and maintain communication with individuals in quarantine and isolation. Healthway employees also supported the general response (email and phone) for questions of all kinds about COVID-19 testing, travel restrictions and BU policies.

#### 1.14 Internal and external dashboards

Data dashboards were developed both for external and internal use.  BU’s [COVID-19 Data Dashboard](https://www.bu.edu/healthway/community-dashboard/) was published to provide transparency and current information on testing of the University community, including daily and multi-day average results by test administered and individuals tested, along with processing time, and local county and state benchmark data.  For internal use, a series of interactive dashboards and reports were developed to monitor and analyze trends in positive cases and their contacts, capacity within the testing centers and laboratories, local and off-campus transmission, and student and employee test and health screening compliance and associated disciplinary action.

#### 1.15 Campus COVID-19 communications

The [Back2BU](https://www.bu.edu/back2bu/student-health-safety/quarantine-isolation-housing/) website provided a robust external communications platform related to the University's COVID-19 response and Q/I housing initiative.^4^ The communications program’s overriding objective was to educate members of the BU community about the disease, its transmission and best risk mitigation practices, and to promote compliance with our health and safety protocols.  The plan rested on five planks:

1. **Provision of** **comprehensive, baseline information** about health and safety protocols and those best practices which enabled BU to operate as a residential academic community during the pandemic.  This information and a resource “bible” were found in the various Back2BU online guidebooks, each updated and customized by cohort (staff, faculty, undergraduate, graduate, etc.)
2. **Daily news streams** were provided to the community with the most up-to-date news and information about the virus, its impact on the community, and changes to health and safety protocols based on new state mandates, discussions with peer institutions, as well as interpretation of data from the internal dashboard and surrounding communities.  BU Today, The Brink and social media accounts were the principal distribution channels for these daily news streams and the new information they contained.
3. **Highly visible public service campaigns** were conducted to elevate awareness of risk and mitigation measures, and influence community members to behave in ways consistent with best health and safety practices. The university’s *Don’t Go Viral* campaign and the student run *F*CKIT* initiative were the delivery mechanisms for that part of the program.
4. **Transparency was used to establish the university as a credible source of information and instruction** pertaining to the pandemic and how to manage it.  The daily dashboard and weekly testing and compliance reports in BU Today, on-line town meetings, along with frequent letters from senior leadership to all segments of the campus community were just a few of the ways BU leadership attempted to be transparent and earn trust and credibility.
5. **Continually monitoring and soliciting feedback** from the community was done to assess the effectiveness of communications and necessary adjustments to make.  Monitoring social media, tracking reader engagement with BU Today, and reports “from the field” provided a good amount of this feedback.

#### 1.16 Enforcement measures

During the course of the semester, a number of different enforcement measures were implemented in order to help reduce transmission of SARS-CoV-2 and to improve compliance among students, faculty, and staff. Student gatherings of all kinds were limited initially to 25 people or less. Given evidence of growing off campus transmission and clusters associated with parties and other gatherings, the Dean of Student’s office reduced the allowable limit of gatherings to 10 people or fewer on November 2, 2020.

To improve adherence to daily health attestations and testing requirements, warning emails for noncompliance were sent from 9/2/2020 to 9/13/2020; however, this was not a confident process due to challenges with precision of the data used to identify noncompliance. On 9/14/2020, official warnings to students and deactivation of student ID card use and WIFI access and banning access to classes began (excluding the medical campus). After two warnings, non-compliant students were prohibited from participating in any classroom or academic activity, either in person or remote (including the ability to access their courses through Blackboard or other platforms, or complete quizzes, examinations, and other course assignments). On 9/21/2020, compliance tracking began on the medical campus, but no warnings or deactivations were issued apart from students at the School of Public Health. On 9/22/20, the remainder of the medical campus was brought into the compliance process. On 9/23/2020, the Dean of Students began managing the compliance database and sending compliance emails to students. On 10/22/2020, badge checking at the entrance of the George Sherman Union began. On 10/27/2020, the process of pulling data from Healthway changed to more accurately assess compliance. The COVID-19 Compliance Manager was brought into the Dean of Student’s office on 11/3/2020. A paradigm shift in corrective action was implemented on 11/9/20 with compliance effort alignment with the University’s Learn from Anywhere indicator to more directly track student location and basis for compliance tracking. In addition, second and third warning emails were eliminated with only one warning followed by deactivation 48 hours later.

#### 1.17 Safe behavior quality improvement project

More than 8,000 safe behavior observations were recorded from August to December 2020. Students were trained to record instances of safe behavior (i.e., wearing masks properly, maintaining social distance after 15 minutes of close contact, and sanitizing hands after touching a new surface) with a standardized Qualtrics-based checklist that was sent to their cell phones.  After completing training and a field test with the manager of the behavioral surveillance team during which students and the Behavioral Surveillance Manager had to achieve 90% inter-rater agreement, students were dispatched to several areas of the campus and completed observational periods of 30 minutes or 30 total observations which ever came first.

During safety observations, student observers noted that lower rates of safe behavior (defined as wearing masks properly, maintaining social distance after 15 minutes of close contact, and sanitizing hands after touching a new surface) occurred during evenings and weekends.  In response, safety promotion interventions were implemented with activities such as moving hand sanitizers, sharing data about safe behavior rates with managers and posting charts daily and subsequently behavior trends against safety targets improved.

#### 1.18 Fall 2020 move-in

Upon arrival, students followed a managed process that included physical distancing, wearing masks, and visiting one of the testing centers to complete a COVID-19 test on their move-in day. Campus move-in was spread out over three weeks rather than the traditional three days. Move-in appointments were required in the morning or afternoon and only one person could help with the move. On arrival, students had to immediately schedule a test and then quarantine in their room (stay-in-place, including attending classes remotely) other than going out to obtain food or have additional SARS-CoV-2 tests performed. After receiving three negative tests over a 10-day period, they no longer had to stay-in-place. Graduate students followed the same approach although they only had to have two negative tests.

### 2. Figures

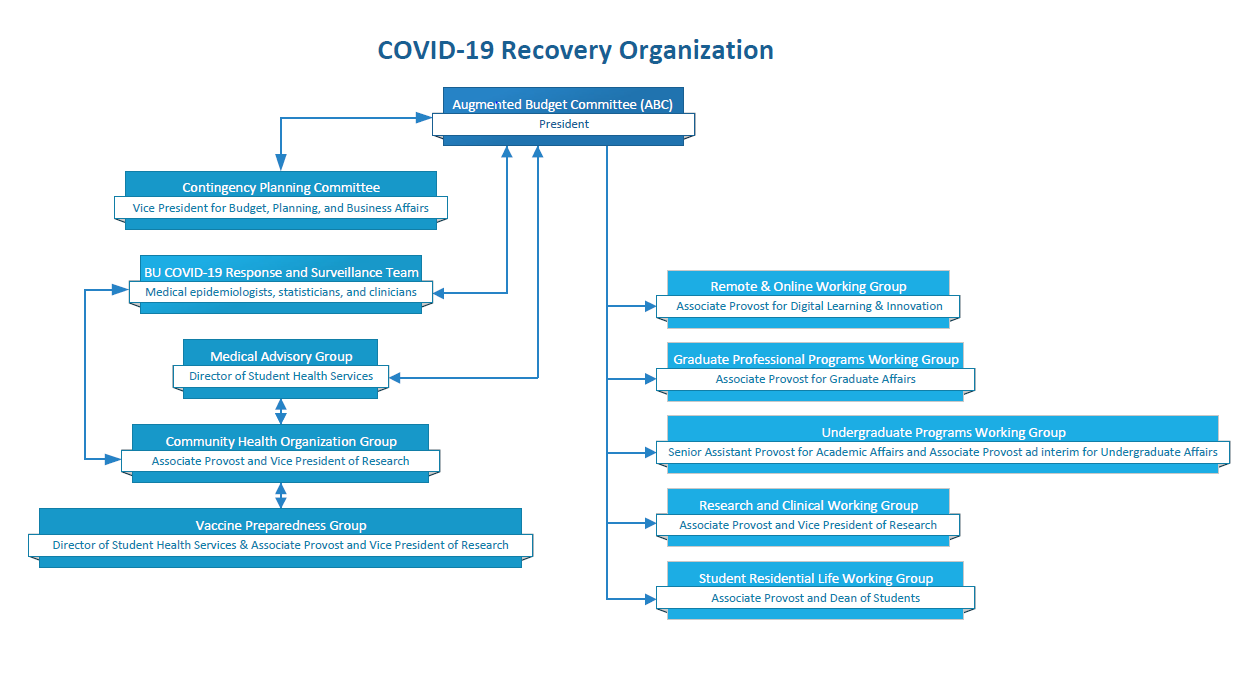

**Figure S1.** Organogram of Boston University’s COVID-19 Recovery Organization

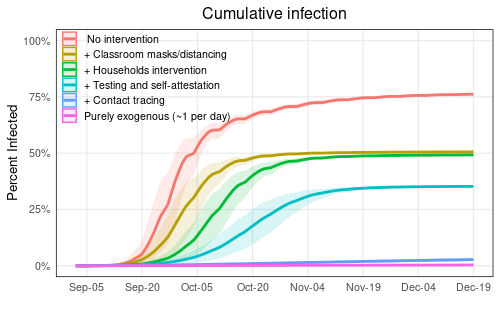

**Figure S2.** Modeling results indicating the relative impact of layering interventions. With no interventions, the model estimated that approximately 75% of the population would be infected and cases would rise exponentially. By incorporating all proposed interventions, this decreased dramatically to a rate reflecting almost no within campus transmission.

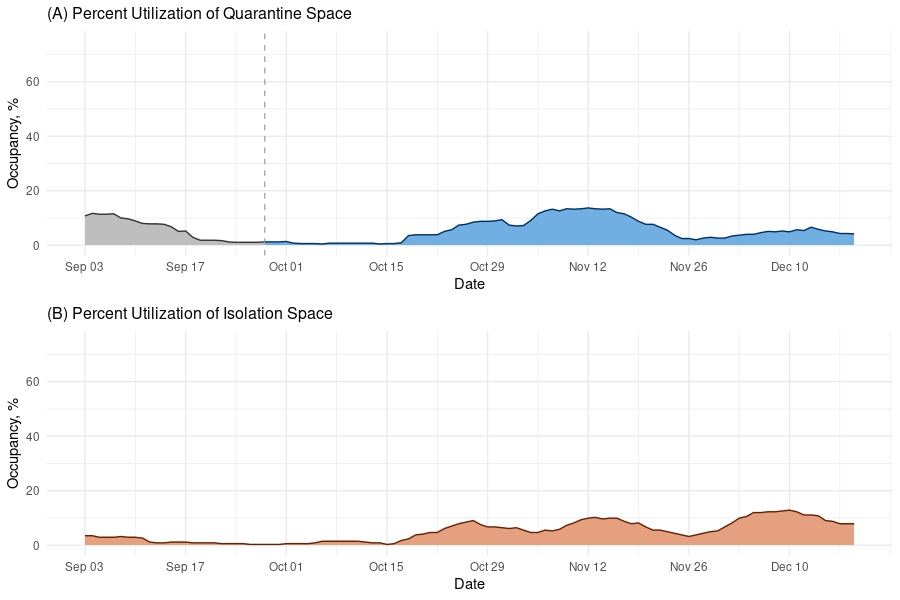

**Figure S3**. Percent utilization of A) quarantine and B) isolation housing throughout the semester. Quarantine data up to September 28 were not collected in a standard format, leading to irregularities.
